# Pre-Exposure Prophylaxis with Various Doses of Hdroxychloroquine among high-risk COVID 19 Healthcare Personnel: CHEER randomized controlled trial

**DOI:** 10.1101/2021.05.17.21257012

**Authors:** Fibhaa Syed, Mohammad Ali Arif, Rauf Niazi, Jaffer Bin Baqar, Ume Laila Hashmi, Sadia Batool, Sadia Ashraf, Junaid Arshad, Saira Musarrat

## Abstract

**Background:** Pre-exposure prophylaxis (PrEP) is a promising strategy to break the chain of transmission of novel coronavirus (2019-nCoV).

**Aims:** This trial aimed to evaluate the safety and efficacy of PrEP with various doses of HCQ against a placebo among high-risk healthcare providers (HCPs).

**Methods:** A phase II, randomized, placebo-controlled trial was conducted at a tertiary care hospital. A total of 228 HCPs were screened, we included 200 subjects with no active or past SARS-CoV-2 infection. Subjects of experimental groups 1-3 received HCQ in various doses and those in the control group received placebo. The study outcomes in terms of safety and efficacy were monitored. Participants exhibiting COVID-19 symptoms were tested for SARS-CoV-2 during the study and also by the end of the 12th week, with PCR or IgM and IgG serology.

**Results:** Overall, 146 of 200 participants reported exposure to a confirmed COVID-19 case in the first month, 189 in the 2nd month and 192 were exposed by the 12th week of the study. Moreover, the precautionary practices, i.e. use of personal protective equipment (PPE), significantly varied; initially more than 80% of the exposed HCPs weren’t ensuring the PPE used by the patients treated by them. However, it gradually developed with the increasing knowledge of the virus. As far as safety is concerned, mild treatment-related side effects were observed among the interventional and placebo arm patients. While none of the participants were critical, and a few had mild illness by the end of the 12th week, requiring only outpatient observation with no hospitalization. There was no significant clinical benefit of PrEP with HCQ as compared to placebo (p>0.05).

**Conclusion:** It is concluded from the study findings that the PrEP HCQ does not significantly prevent illness compatible with COVID-19 or confirmed infection among high-risk HCPs.

## Introduction

Novel severe acute respiratory syndrome coronavirus 2 (SARS-CoV-2) has by far affected almost all countries. Globally as of February 24, 2021, there have been about 111,593,583 confirmed cases of COVID-19 and 2,475,020 deaths, as per World Health Organization (WHO) emergency statistics [1]. For the proper management of this pandemic, the most crucial step is the conservation of workforce safety. As this outbreak holds a heavy toll on the frontline healthcare workers, they are three times more likely to be infected than unexposed [2]. Moreover, this pandemic has affected us physically and psychologically; the exposed frontline workers frequently developed depressive symptoms, anxiety, fear, and stress [3-5].

Given the healthcare workers exposed to COVID-19 patients are at high risk of viral transmission. Therefore, many of them stayed away from their homes for keeping their families safe [6]. The fight against COVID-19 is in continuance and the success only relies on the adherence to the preventive measures. A series of preventive measures and therapeutic options are being explored for disease containment. In addition to the quarantine of exposed individuals from close ones, the prevention strategies also include the use of personal protection equipment (PPE), hand washing, case identification and isolation [7-9]. Numerous drugs are undergoing clinical trials for effective mitigation of SARS-CoV-2 transmission [6,10]. Until now, remdesivir is the only drug approved by the Food and Drug Administration (FDA) to treat COVID-198. Besides, dexamethasone also improved the disease outcomes among severe COVID-19 patients [11,12].

In addition, research shows that chloroquine acts as an effective in vitro inhibitor of SARS-CoV-2. Furthermore, HCQ, a chloroquine derivate is also identified as a possible prophylactic inhibitor for the entry and post-entry stages of SARS-CoV-2, with better in vitro antiviral activity and safety profile, on the ground of anti-inflammatory as well as antiviral effects [13-15]. HCQ has been evaluated for the treatment of SARS-CoV-2 (pneumonia) and post-exposure prophylaxis. The available clinical evidence for the use of HCQ among COVID-19 patients seems insufficient and does not fully prove the effectiveness of this therapeutic modality among severely ill COVID-19 patients. Nevertheless, the promising treatment outcomes observed among mildly infected patients and no environmental implications associated with the drug suggest its therapeutic potential.

Pre-exposure Prophylaxis (PrEP) is suggested to be a promising strategy for several infectious diseases [16], but none of the pre-exposure pharmacological drugs have been established for COVID-19 yet. However, this study aims to evaluate the therapeutic safety and efficacy of PrEP with various doses of HCQ among the healthcare workers who are at high risk for COVID-19 disease in the period of this epidemic.

## Materials and Methods

### Study design & participants

A Phase II, randomized, placebo-controlled clinical trial (Clinicaltrials.gov NCT04359537) was conducted at SZABMU/PIMS to evaluate the comparative efficacy of various HCQ doses in preventing COVID-19 among high-risk healthcare workers. Enrolment began on 1st May 2020, and the intervention continued for a total of 12 weeks.

All healthcare workers at high risk for COVID-19 exposure, primarily those in emergency departments (physicians, nurses, ancillary staff, triage personnel), intensive care units (physicians, nurses, ancillary staff, respiratory therapists), performing aerosol-generating procedures (anesthesiologists, nurse, anesthetists, gastroenterologists performing endoscopy, pulmonologists performing bronchoscopy), first responders (EMTs, paramedics) and those working in the departments of general medicine, pulmonology, infectious disease and isolation wards were included in the study. While active COVID-19 cases, those with existing symptoms like fever, cough, shortness of breath, having prior retinal eye disease, Chronic Kidney Disease (CKD), Stage 4 or 5 or dialysis, Glucose-6 Phosphate Dehydrogenase (G-6-PD) deficiency, recent Myocardial Infarction (MI) and epileptic subjects were excluded. Additionally, also kept under exclusion were pregnant females, subjects weighing < 40 kg, those having contraindication or allergy to chloroquine/ HCQ, already administering HCQ or cardiac medicines like flecainide, amiodarone, digoxin, procainamide, or propafenone, medications with known significant drug-drug interactions like artemether, lumefantrine, mefloquine, tamoxifen or methotrexate and those causing QT interval prolongation like macrolides, antipsychotics, quinolones, antihistamines, SSRIs, tricyclic antidepressants, antifungals.

A total of 228 participants were initially enrolled (Figure 1); of them, 28 were ineligible and excluded. Participants fulfilling the eligibility criteria were randomized into the four treatment groups. Group 1 participants (n=48) were intervened with HCQ 400 mg (locally manufactured by Getz Pharma) twice a day on day 1 followed by 400 mg weekly. Group 2 (n=51) participants were given HCQ 400 mg once every 3 weeks, group 3 (n=55) administered HCQ 200 mg once every 3 weeks and participants in the control group received placebo (n=46).

**Figure 1:**
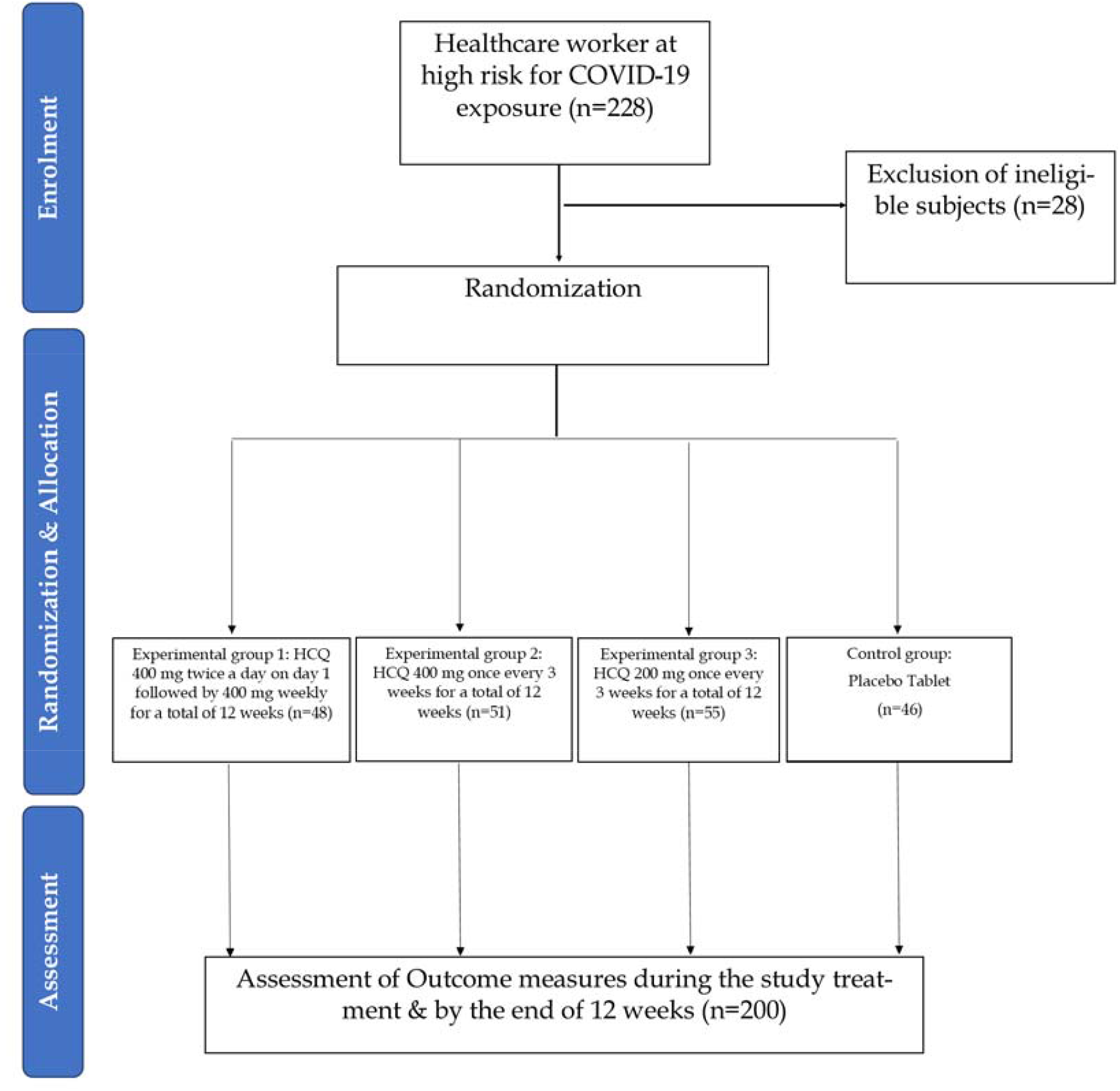
Flowchart of the study.

### Assessments & Follow-up Procedure

The baseline characteristics of all participants, including age, gender, role, comorbidities, and drug records, were obtained. COVID-19 related symptoms and adverse events (AEs) from the drug were self-reported by the enrolled participant during the study period. The COVID-19 exposure and preventive practices were monitored on a monthly basis. Disease severity was assessed through ordinal scale i.e. no illness (score=1), illness with outpatient observation (score=2), hospitalization (or post-hospital discharge) (score=3), hospitalization with ICU stay (score=4) and death from COVID 19 (score=5). All participants exhibiting COVID-19 symptoms were tested for SARS-CoV-2 during the study and also by the end of the 12th week, with PCR or IgM and IgG serology (as per accessibility).

### Outcomes

The primary endpoint was to evaluate the COVID-19-free survival among the participants by the end of the study. The secondary endpoints were to evaluate the proportion of rRT-PCR positive COVID-19 cases, the role of exposure and preventive practices, the frequency of COVID-related symptoms, treatment-related side effects, the incidence of all-cause study medicine discontinuation, and maximum disease severity during the study treatment.

### Statistical Methods

The continuous variables were summarized as means and standard deviations (SD) and categorical variables as frequencies and percentages. A comparative analysis was performed between the experimental groups. Chi-square test and analysis of variance (ANOVA) were used to compare the baseline characteristics, COVID-19 exposure, preventive measures, symptomatology, side effects and maximum disease severity. A p-value <0.05 was considered significant.

### Ethical Consideration

The study protocol was approved by the ethical review board of Shaheed Zulfiqar Ali Bhutto Medical University (Reference no. 1-1/2015/ERB/SZABMU/549; Dated 20 April 2020), and written informed consents were acquired from the participants before inclusion.

### Results

The baseline characteristics of the enrolled healthcare personnel are shown in table 1. The trial included 109 (54.5%) male participants, and the mean age was 30.63 ± 8.075 years. The majority of them were doctors and had no comorbid conditions.

**Table 1:** Demographic characteristics of the studied population (n=200)

| Variable |  | Experimental Group 1 (n=48) | Experimental Group 2 (n=51) | Experimental Group 3 (n=55) | Control Group (n=46) | p-value |
| --- | --- | --- | --- | --- | --- | --- |
| Age (years); Mean ± SD | | 30.40±7.764 | 28.20±4.486 | 32.02±9.517 | 31.91±9.13 | 0.057 |
| Gender | Male | 24(50) | 25(49.0) | 37(67.3) | 23(50) | 0.172 |
|  | Female | 24(50) | 26(51.0) | 18(32.7) | 23(50) |  |
| Profession | Doctor | 32(66.7) | 43(84.3) | 43(78.2) | 25(54.3) | 0.050 |
|  | Nurse | 9(18.8) | 4(7.8) | 7(12.7) | 9(19.6) |  |
|  | Technician | 1(2.1) | - | 2(3.6) | 5(10.9) |  |
|  | Emergency Staff | 1(2.1) | - | 1(1.8) | - |  |
|  | Emergency Responder | - | 2(3.9) | - | - |  |
|  | Sanitary Worker | 2(4.2) | 1(2.0) | - | 2(4.3) |  |
|  | Security Guard | 3(6.3) | 1(2.0) | 2(3.6) | 5(10.9) |  |
| Comorbid Conditions | None | 45(93.8) | 50(98.0) | 48(87.2) | 41(89.1) | 0.222 |
|  | Diabetes | 2(4.2) | - | 2(3.63) | 3(6.5) |  |
|  | Hypertension | 1(2.1) | 1(2.0) | 5(9.09) | 2(4.3) |  |
| Department | Medicine | 23(47.9) | 29(56.9) | 23(41.8) | 28(60.9) | 0.296 |
|  | Pulmonology | 3(6.3) | 1(2.0) | 2(3.6) | 1(2.2) |  |
|  | ICU | 3(6.3) | 5(9.8) | 5(9.1) | 3(6.5) |  |
|  | Emergency | 2(4.2) | 4(7.8) | 4(7.3) | 2(4.3) |  |
|  | Gastroenterology | 1(2.1) | 1(2.0) | - | 1(2.2) |  |
|  | Nephrology | 1(2.1) | - | - | 5(10.9) |  |
|  | Gynae & Obs | 3(6.3) | 2(3.9) | 4(7.3) | 1(2.2) |  |
|  | Pediatric | 4(8.3) | 1(2.0) | 4(7.3) | 1(2.2) |  |
|  | Surgery | 3(6.3) | 4(7.8) | 3(5.5) | - |  |
|  | ENT | - | 1(2.0) | 3(5.5) | 2(4.3) |  |
|  | Radiology | - | 1(2.0) | - | 1(2.2) |  |
|  | Pathology | 1(2.1) | - | 3(5.5) | - |  |
|  | Neurology | 1(2.1) | 1(2.0) | 3(5.5) | - |  |
|  | Cardiology | 3(6.3) | 1(2.0) | 1(1.8) | 1(2.2) |  |
| Smoking Status | Smoker | 5(10.4) | 7(13.7) | 7(12.7) | 7(15.2) | 0.917 |
|  | Non-Smoker | 43(89.6) | 44(86.3) | 48(87.3) | 39(84.8) |  |

\*Values are given as n(%) or Mean ± SD
\*Experimental Group 1-HCQ (400 mg BD on day 1 followed by 400 mg once weekly); Experimental Group 2-HCQ (400 mg once every 3 weeks); Experimental Group 3-HCQ (200 mg once every 3 weeks); Control Group-Placebo

There was no significant difference in the exposure records between the treatment groups as shown in table 2. Most of these healthcare workers were exposed to a confirmed or probable COVID-19 case. Around 73% of participants were exposed in the first month (baseline), which further increased to 94.5% by the second month (1^st^ follow-up) and 96% by the end of the 12^th^ week (2^nd^ follow-up). Initially, 18.5% of participants weren’t using PPE, but with the passage of time, the awareness regarding preparedness developed and by the end of the 12^th^ week, only 4.5% of participants weren’t taking precautionary measures.

**Table 2:**
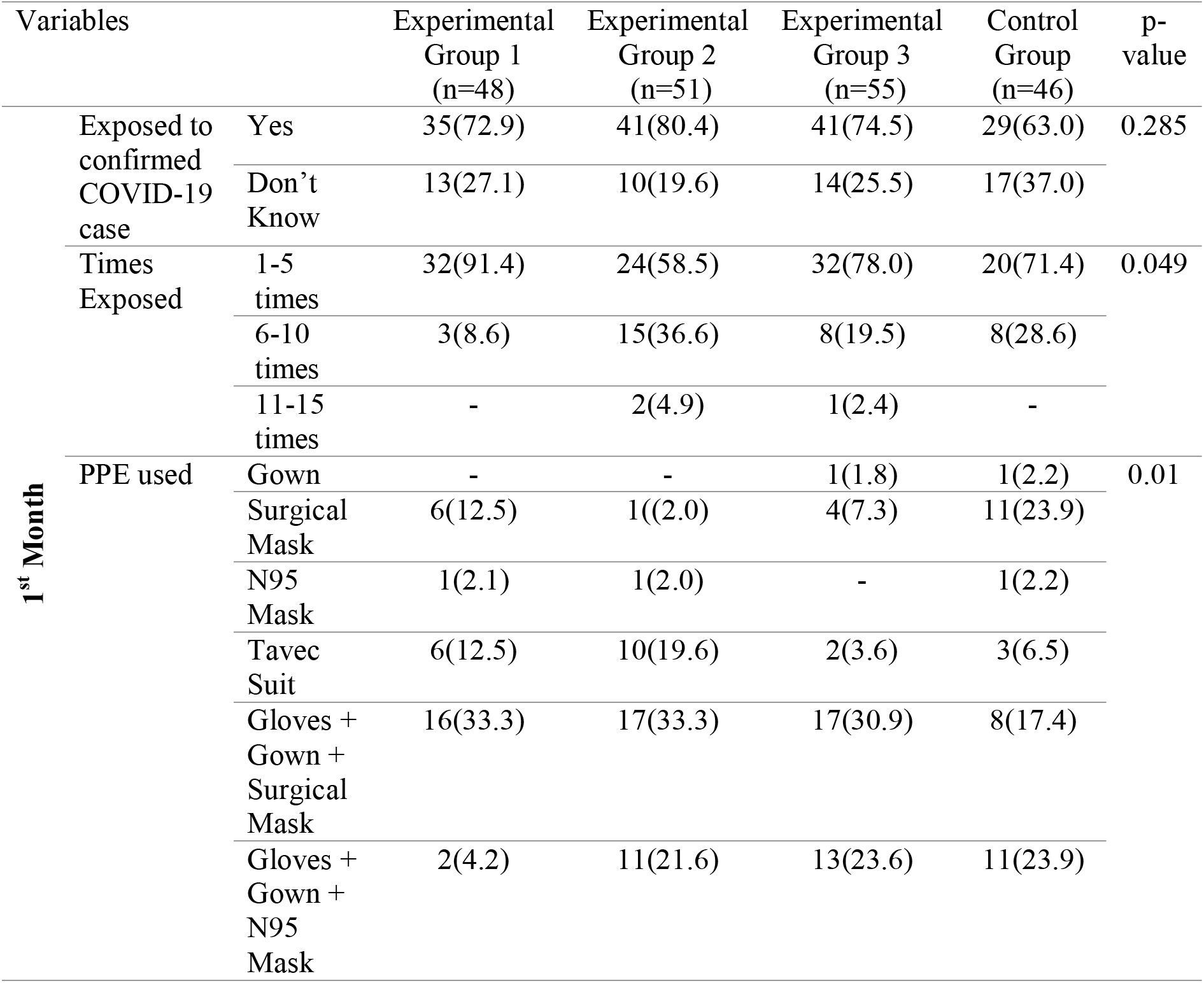
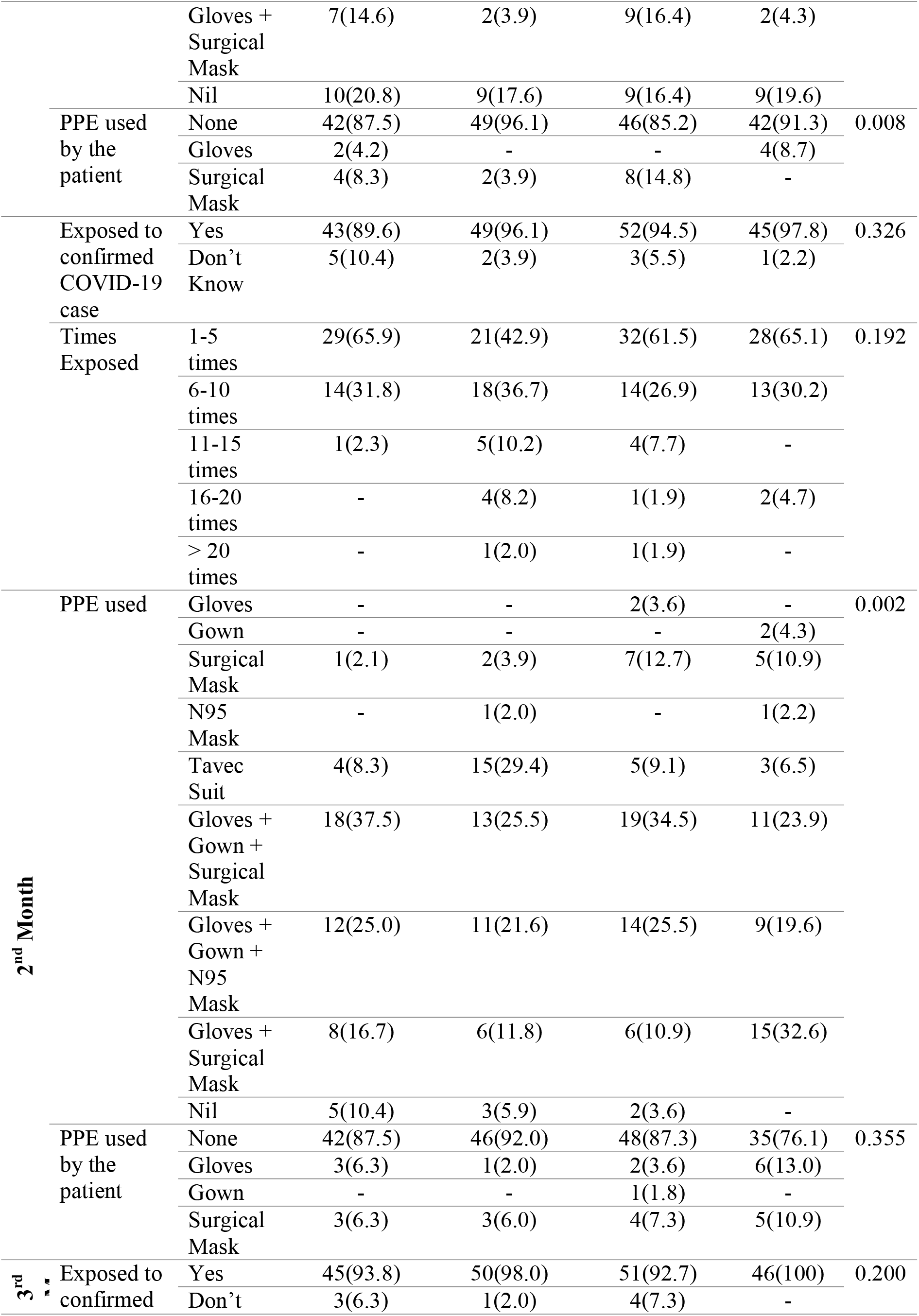
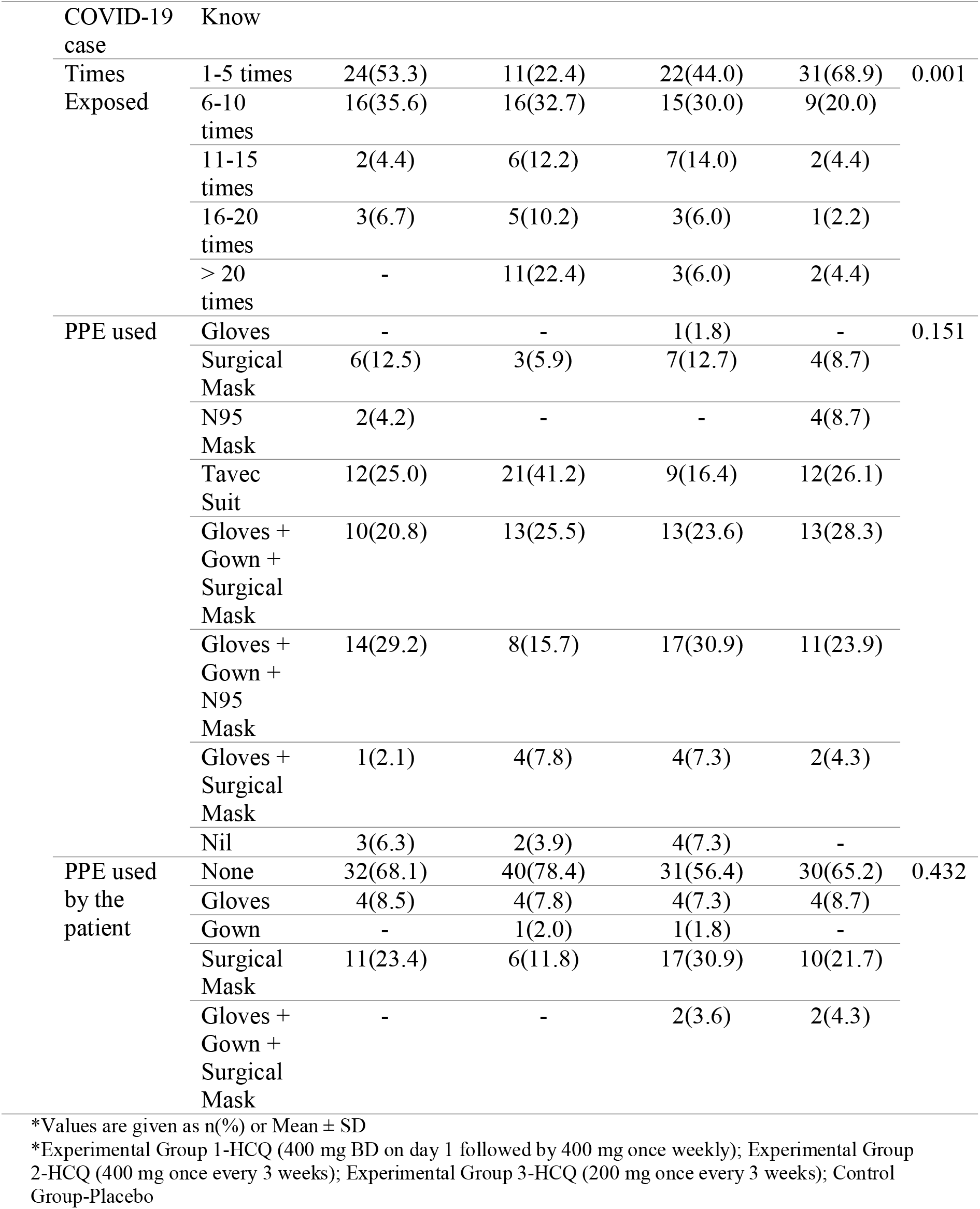
Monthly evaluation of COVID exposure & prevention.

Of the total, COVID-19 related symptoms appeared in 54.9% participants of experimental group 2, 33.3% of group 1, 30.4% from the placebo group and 23.6% from group 3 (Table 3). Fever, cough, shortness of breath, rhinorrhea, and diarrhea were the commonly reported symptoms. The frequency of cough was significant among the participants of experimental group 2 receiving HCQ (400 mg once every 3 weeks).

**Table 3:** COVID related symptoms.

| Variables |  | Experimental<br>Group 1<br>(n=48) | Experimental<br>Group 2<br>(n=51) | Experimental<br>Group 3<br>(n=55) | Control<br>Group<br>(n=46) | p-value |
| --- | --- | --- | --- | --- | --- | --- |
| Symptomatology | Yes | 16(33.3) | 28(54.9) | 13(23.6) | 14(30.4) | 0.006 |
|  | No | 32(66.7) | 23(45.1) | 42(76.4) | 32(69.6) |  |
| Fever | No | 31(64.6) | 25(49.0) | 42(76.4) | 33(71.7) | 0.037 |
|  | Mild | 15(31.3) | 17(33.3) | 9(16.4) | 8(17.4) |  |
|  | Moderate | 2(4.2) | 9(17.6) | 4(7.3) | 5(10.9) |  |
| Cough | No | 32(66.7) | 31(60.8) | 46(83.6) | 33(71.7) | 0.057 |
|  | Mild | 11(22.9) | 18(35.3) | 6(10.9) | 12(26.1) |  |
|  | Moderate | 5(10.4) | 2(3.9) | 3(5.5) | 1(2.2) |  |
| Shortness of<br>breath | No | 43(89.6) | 49(96.1) | 53(96.4) | 44(95.7) | 0.271 |
|  | Mild | 5(10.4) | 1(2.0) | 1(1.8) | 2(4.3) |  |
|  | Moderate | - | 1(2.0) | - | - |  |
|  | Severe<br>(Requiring<br>Hospitalization) | - | - | 1(1.8) | - |  |
| Rhinorrhea | No | 33(68.8) | 33(64.7) | 51(92.7) | 38(82.6) | 0.002 |
|  | Mild | 13(27.1) | 15(29.4) | 4(7.3) | 8(17.4) |  |
|  | Moderate | - | 3(5.9) | - | - |  |
|  | Severe<br>(Requiring<br>Hospitalization) | 2(4.2) | - | - | - |  |
| Diarrhea | No | 45(93.8) | 51(100) | 53(96.4) | 43(93.5) | 0.316 |
|  | Mild | 3(6.3) | - | 2(3.6) | 3(6.5) |  |

\*Values are given as n(%) or Mean ± SD
\*Experimental Group 1-HCQ (400 mg BD on day 1 followed by 400 mg once weekly); Experimental Group 2-HCQ (400 mg once every 3 weeks); Experimental Group 3-HCQ (200 mg once every 3 weeks); Control Group-Placebo

No serious AEs were observed. Five participants from experimental group 1, one from experimental group 2, three from experimental group 3 and one from the control group reported treatment-related side effects. During the treatment, 15 out of 48 in group 1, 19 out of 51 in group 2, 8 out of 55 in group 3, and 7 out of 46 in control group tested positive for COVID-19. More than 70% of the studied participants (from all studied groups) displayed no illness. While 27.5% of group 2 exhibited illness (outpatient observation). The COVID-19 test results were similar among the treatment groups by the end of 12 weeks; the PCR results were negative among almost all the study participants, while 6.5% of participants from the placebo arm tested positive. The samples were also tested for serology, results shown in table 4.

**Table 4:**
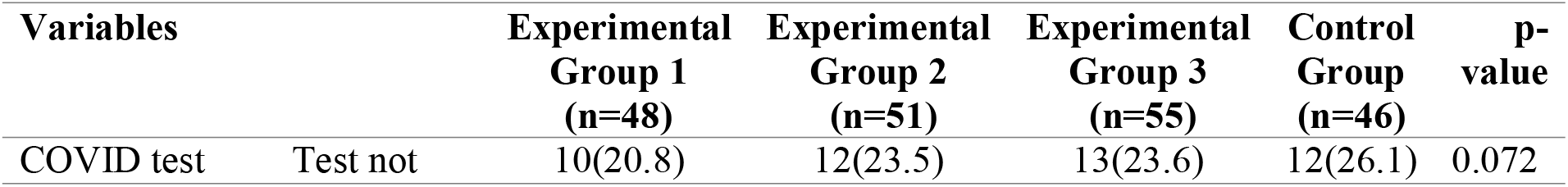
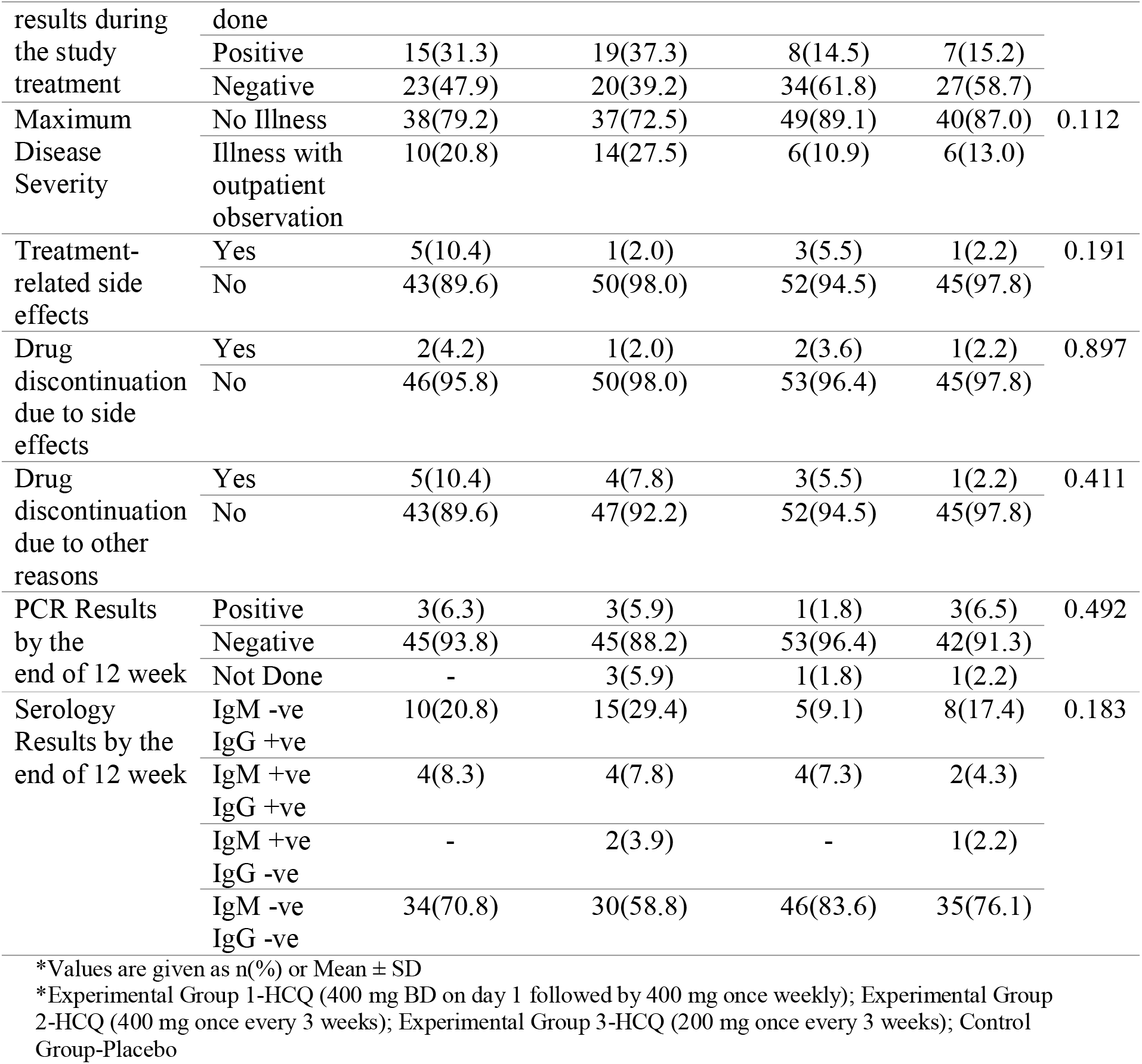
Relationship of management and laboratory test profile of trial subjects.

## Discussion

A Phase II, randomized, placebo-controlled clinical trial was conducted to evaluate the effectiveness of various doses of HCQ in PrEP for the prevention of COVID-19 among high-risk healthcare workers. As they are at the highest risk of COVID-19 infection, it is essential to ensure adequate allocation of PPE to alleviate the structural inequities associated with COVID-19 risk. The results of the present study suggest no significant difference in the exposure records between the treatment groups. The rate of exposure to a confirmed COVID case increased by the end of the 12th week (2nd follow-up) among the participants of all treatment groups. Although the rate of involvement in the preventive practices wasn’t very promising initially, but with progressing knowledge and awareness, the level of preparedness excelled among the healthcare personnel of all study groups. Our finding suggests a consistency with a similar randomized trial related to SARS-CoV-2 and PrEP, with the dosage of HCQ [17]. Moreover, other similar observational studies also suggest an increased risk with the inadequate use of PPE among healthcare workers. It is to note that even with adequate PPE use and preventive practices, the healthcare personnel remain on the maximum COVID-19 risk [18].

During the first month of the study, the overall accumulated incidence of COVID-19 among the study participants was 32.02%. The rate of COVID-19 positivity was similar in the HCQ and placebo arms (p=0.072). A subsequent decrease in the COVID-19 incidence was observed by the PrEP with HCQ by the end of the 12th week, as per the PCR results. Moreover, the infection rate was lowest among the participants treated with the low drug dose (200 mg once every 3 weeks). Grau-Pujol et al., in their study, concluded that PrEP with low doses of HCQ is safe and effective [19]. Furthermore, most of our study participants (83.6%) tested negative for IgG and IgM antibodies by the end of 12 weeks. In support, a randomized clinical trial also reported a similar COVID-19 incidence rate among the participants of HCQ and placebo arms, i.e. 6.3% vs. 6.6% (p > 0.99) [20]. Likewise, Boulware et al. suggested no significant difference in the incidence of COVID-19 after the treatment with HCQ than those given placebo only, with additional gastrointestinal and neurologic side effects among the participants of interventional arm [17].

In this trial, the types and frequency of symptoms reported in the present study were similar to those in previous studies involving HCPs [21,22]. It was also observed that the treatment-related side effects were comparatively more evident among the participants of the interventional arms than placebo, i.e. 10.4%, 2.0% and 5.5% participants from experimental group 1, 2 and 3, respectively, receiving various doses of HCQ vs. 2.2% of the placebo group were observed with treatment-related side effects. However, none of the participants required hospitalization, ICU care or death from COVID-19 in any study groups. A majority reported no illness in response to the provided treatment, and a few had a mild illness (outpatient observation). Our safety data is similar to a randomized clinical trial indicating a higher rate of adverse events reported among the intervention arm patients than those receiving placebo [23]. Moreover, five participants discontinued the HCQ prophylaxis due to side effects and one in the placebo arm; these findings are consistent with a similar study [23].

Adding to the existing literature, this trial provided a detailed analysis of the monthly exposure history, intensity of exposures, and the enrolled healthcare personnel’s preventive practices. However, we acknowledge the limitations. The small sample size for assessing the efficacy of PrEP with HCQ at the initial stages of analysis among the healthcare workers was one of the major limitations. Based on the findings, the incidence rate of SAR-CoV-2 in healthcare workers declined from the initiation of the study till the end of the analysis. Moreover, it cannot be denied that a lower HCQ dose effectively prevented the COVID-19 infection in the studied cohort. Further large-scale prophylaxis trials are required to investigate the antiviral activity of varying HCQ dosing and the differential impact of each therapeutic agent on the body’s biochemical profile and the overall disease incidence.

## Conclusions

In conclusion, there was no significant reduction in the SARS-CoV-2 transmission with PrEP administration of Hydrochloroquine among the enrolled healthcare personnel. The safety profile varied among the participants who were intervened with HCQ compared to those given placebo, the presence of treatment related side-effects was higher among the participants of experimental group 1 and 3 receiving different HCQ doses. In contrast, the safety outcome of experimental group 2 was similar to that observed in the placebo arm. As far as the disease severity is concerned, none of the participants were severe/critical enough to require hospitalization and ICU care, and none of them died. Preventing healthcare workers from COVID-19 is critical to control the pandemic. Thus, further large-scale clinical studies are required to investigate the safety and efficacy of the various doses of HCQ in order to break the inciting viral transmission.

## Data Availability

The data supporting the study conclusions is available at Figshare.

https://figshare.com/articles/dataset/Pre-Exposure_Prophylaxis_with_Various_Doses_of_Hdroxychloroquine_among_high-risk_COVID_19_Healthcare_Personnel_CHEER_randomized_controlled_trial_/14572527

## Data Availability

The data supporting the study conclusions is available at Figshare (https://figshare.com/articles/dataset/Pre-Exposure_Prophylaxis_with_Various_Doses_of_Hdroxychloroquine_among_high-risk_COVID_19_Healthcare_Personnel_CHEER_randomized_controlled_trial_/14572527)

## Acknowledgments

The authors would like to acknowledge the Medical Affairs department of Getz Pharma for their technical support and assistance in the publication process.

